# A super-series of extracellular miRNA in cerebrospinal fluid reveals broad patterns associated with neurologic disease

**DOI:** 10.1101/2020.05.20.20108472

**Authors:** Andrew Dhawan

**Affiliations:** Department of Neurology, Neurological Institute, Cleveland Clinic Foundation, Cleveland, OH, USA, 44195

**Keywords:** Biomarker, non-coding RNA, CSF, miRNA, neurologic disease

## Abstract

Extracellular non-coding RNA are emerging biomarkers across diseases, as they may represent the functional state of the cells of origin, and have been robustly detected and associated with disease in many biofluids, but not yet at a large scale in cerebrospinal fluid (CSF). While there have been extracellular non-coding RNA (exmiRNA) identified in small associative studies involving the CSF and patients with neurologic conditions, much remains to be determined, including what constitutes healthy CSF. In this work, we aggregate 9 existing cohorts to define a super-series of 864 extracellular circulating microRNA (exmiRNA) samples in the CSF from patients with 16 neurologic diseases and healthy controls. Through this, we identify broad patterns of conserved expression of CSF exmiRNA in the healthy state and in neurologic diseases, and a set of robustly detectable miRNA that may be strong biomarker candidates for future studies. We show that the coexpression network of exmiRNA differs significantly between the malignant and non-malignant states, suggesting that widespread dysregulation of miRNA due to cancer can be detected in the CSF. The super series will serve as a resource for future research in CSF exncRNA biomarkers, and will continue to evolve as more data is generated in this burgeoning field.

## Introduction

Across the spectrum of neurologic disease, the need for non-invasive biomarkers of illness is massive, given that many conditions are diagnosed based on clinical criteria alone, have significant overlap with other syndromes, and cannot be differentiated until late in the disease course, after irreversible damage has occurred.^1,2^ Moreover, in neurodegenerative conditions with heterogeneous clinical phenotypes, such as Alzheimer’s disease, amyotrophic lateral sclerosis (ALS), or Parkinson’s disease, clinical trials may be limited by difficult identification of optimal candidates for therapies because of difficulties in identifying minimally symptomatic disease, and identifying an appropriate target population.^3–6^ In response to this, there has been a significant effort to identify non-invasive biomarkers of neurologic disease involving combinations of clinical features, radiomic studies, and biochemical testing of the serum and cerebrospinal fluid (CSF).^7–9^

Among these non-invasive biomarkers, testing for extracellular genetic material in the form of exosomes or free-floating fragments of DNA and RNA has emerged as a promising modality. Used particularly in serum testing, as a ‘liquid biopsy,’ these are becoming standard clinical practice in the non-invasive monitoring of solid tumours, and in prenatal diagnostics.^10–12^ Extracellular microRNA (exmiRNA), small 18-22 nucleotide regulatory RNA molecules, have recently been shown to be stable and robustly detectable in nearly every biofluid, including serum, saliva, urine, and cerebrospinal fluid (CSF).^13^ These small molecules act at the transcriptional level to induce the degradation of specific target messenger RNA (mRNA) molecules, ultimately reducing or stabilizing protein levels.^14^ Small variations in their expression, amplified by the pleiotropy of their cognate mRNA targets, result in significant phenotypic changes in the expressing cell.^15,16^ Gaining an understanding of exmiRNA may allow for an *in vivo* functional biomarker of neural cell activity, and may not only inform the diagnosis of disease, but also may allow for a better understanding of disease pathophysiology.

Despite their significant promise, exmiRNA CSF biomarkers have yet to reach mainstream clinical use owing to a multitude of factors. There is limited data in best practices for small RNA extraction from CSF, as well as a limited understanding of the range of expression in healthy patients, and even less of an understanding of the differences in exmiRNA expression across the compendium of neurologic disease.^17^ Larger studies are needed, in concert with the integration of extant data to guide clinicians and researchers to the optimal biomarker candidates.

As a step towards addressing these issues, in this work, primary data from human samples of CSF exmiRNA was aggregated, contacting authors and data repositories for data not publicly available. Datasets were annotated with as much metadata as possible, results have been stored in a harmonized format, and have been made publicly available. Using this super-series of 864 patients across 16 neurological conditions from 9 studies, we studied the impact of technical protocol on exmiRNA yield, and uncovered an understanding of candidate CSF exmiRNA biomarkers across a spectrum of neurological disease. In doing so, this work represents the largest super-series of such patients to date, and identifies candidate biomarkers as well as a set of 30 consensus exmiRNA robustly detected across samples, with 8 species detected strongly in healthy tissues. It is our hope that these findings can be used to guide future work in the development of CSF exmiRNA biomarkers.

## Results

### Assembling a super-series of CSF exmiRNA samples

We sought to identify all possible published and unpublished works of CSF samples with unbiased sequencing of extracellular miRNA, first with a structured literature search in Ovid EMBASE, as described in Methods. Our search strategy returned 83 studies, manually screened for deduplication, primary data, human samples, and unbiased miRNA characterization. In addition to the structured portion of the literature search, an unstructured approach was taken analysing Google Scholar search results for CSF miRNA studies, which yielded an additional 12 studies. Two series (Jensen_IVH, Jensen_SAH) were added from the exRNA database without associated publication, and another was identified directly on the GEO database (Dong et al., 2018, GSE108398).^13^ Datasets were not included if they solely reported relative changes by qtPCR due to the limitations in measuring an unbiased set of exmiRNAs with this method. After screening, 20 studies were identified with likely suitable data, and for those studies in which data was not publicly available, authors were contacted multiple times to report data. Flow diagram of this process is presented in Figure 1. Ultimately, 9 studies had sufficient data to be included in the super-series, as summarised in Table 1.

**Figure 1.**
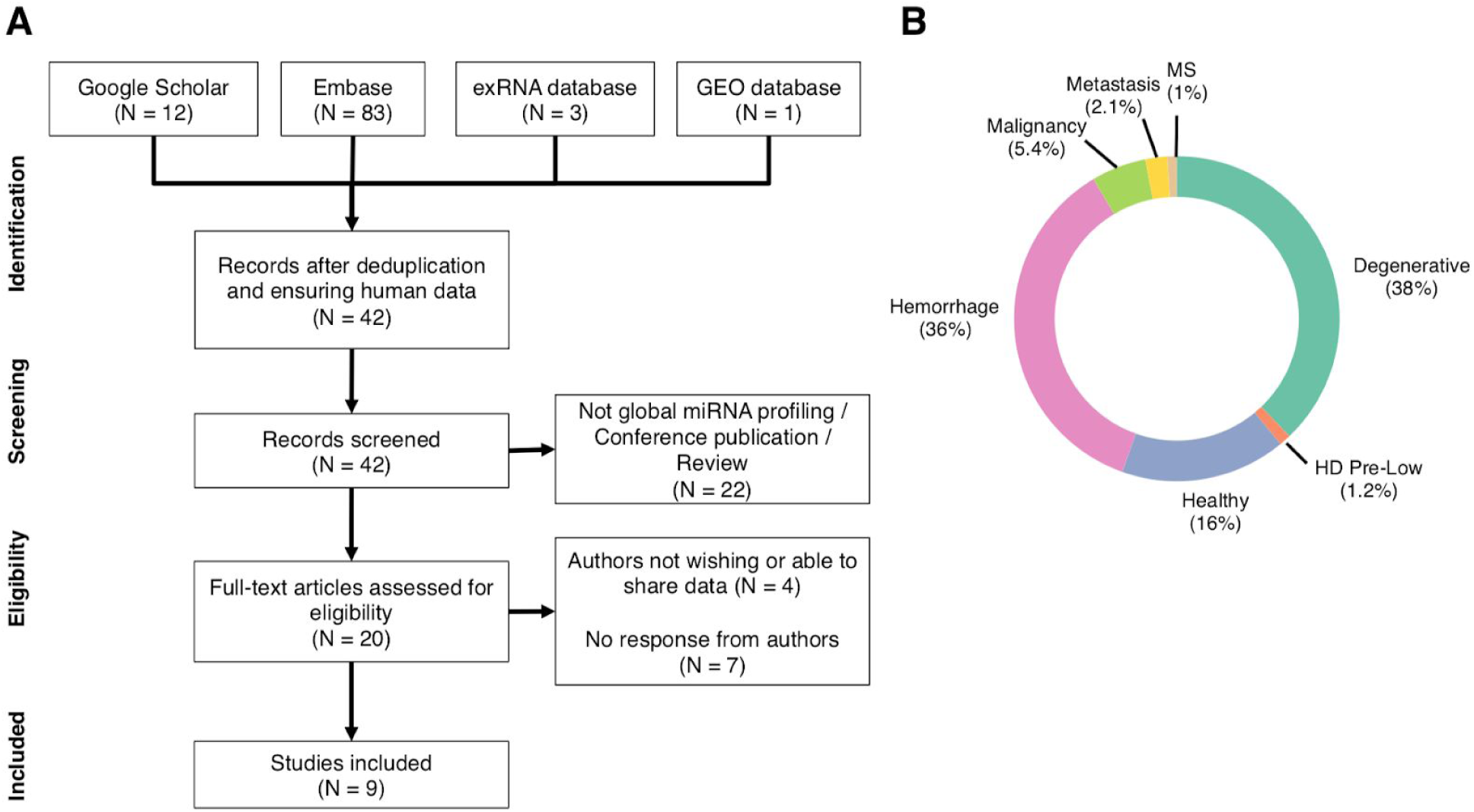
Literature search overview and sample breakdown. (a) PRISMA-style diagram depicting the inclusion and exclusion of relevant studies used to define super-series of CSF exmiRNA samples. (b) Breakdown of patient samples included in super series by proportion. HD Pre-Low refers to the low clinical risk group for Huntington’s disease, as defined by Reed et al. in the PREDICT-HD study ^18^. MS refers to multiple sclerosis.

**Table 1.** Super-series composition. Listing of studies comprising the super-series, as well as patient breakdown by disease, gender, and age. AD refers to Alzheimer’s disease. PD refers to Parkinson’s disease. PDD refers to Parkinson’s disease dementia. M refers to male, F to female. HD refers to Huntington’s disease. GBM refers to glioblastoma. Met refers to metastasis. IVH refers to intraventricular hemorrhage. SAH refers to subarachnoid hemorrhage. HD Pre-Low refers to low clinical risk score for Huntington’s disease, Pre-Med refers to medium clinical risk score for Huntington’s disease, and Pre-High refers to high clinical risk score for Huntington’s disease all as defined in the PREDICT-HD study^18^. For the purposes of this analysis, Pre-Med and Pre-High risk groups were joined with the overall HD group and Pre-low group was kept separate. ALS refers to amyotrophic lateral sclerosis. MS refers to multiple sclerosis.

| Study | Patients | Breakdown | Gender | Age (y) | Reference |
| --- | --- | --- | --- | --- | --- |
| Burgos et al., 2014 | 181 | 62 Healthy<br>62 AD<br>36 PD<br>21 PDD | M: 95 (52%)<br>F: 86 (48%) | 0-19: 0<br>20-39: 1 (0.6%)<br>40-59: 3 (2%)<br>60-79: 77 (43%)<br>80+: 134 (74%) | 19 |
| Dong et al., 2018 | 136 | 21 Healthy<br>115 HD | M: 52 (38%)<br>F: 84 (62%) | 0-19: 0<br>20-39: 59 (43%)<br>40-59: 57 (42%)<br>60-79: 20 (15%)<br>80+: 0 | GSE108398 |
| Drusco et al., 2015 | 80 | 14 Healthy<br>16 GBM<br>15 Medulloblastoma<br>13 Breast Met<br>7 Glioma<br>6 Lung Met<br>4 Meningioma<br>3 Lymphoma<br>2 Ependymoma | Not available | Not available | 20 |
| Godoy et al., 2018 | 8 | 8 Healthy | Not available | Not available | 21 |
| Jensen, IVH | 70 | 70 IVH | M: 51 (73%)<br>F: 19 (27%) | 0-19: 70 (100%)<br>20-39: 0<br>40-59: 0<br>60-79: 0<br>80+: 0 | 13 |
| Jensen, SAH | 241 | 241 SAH | Not available | 0-19: 22 (9%)<br>20-39: 20 (8%)<br>40-59: 140 (58%)<br>60-79: 67 (28%)<br>80+: 8 (3%) | 13 |
| Reed et al., 2018 | 60 | 15 Healthy<br>10 HD-Pre-low<br>35 HD, HD-Pre-Med,<br>HD-Pre-High | M: 27 (45%)<br>F: 33 (55%) | 0-19: 0<br>20-39: 24 (40%)<br>40-59: 24 (40%)<br>60-79: 12 (20%)<br>80+: 0 | 18 |
| Waller et al., 2018 | 82 | 16 Healthy<br>58 ALS<br>8 MS | Not available | Not available | 22 |
| Yagi et al., 2017 | 6 | 6 Healthy | M: 2 (33%)<br>F: 4 (67%) | 0-19: 0<br>20-39: 2 (33%)<br>40-59: 2 (33%)<br>60-79: 2 (33%)<br>80+: 0 | 23 |
| Totals | 864 | 142 Healthy | M: 227 (50%)<br>F: 226 (50%) | 0-19: 89 (13%)<br>20-39: 103 (15%)<br>40-59: 222 (32%)<br>60-79: 161 (23%)<br>80+: 119 (17%) |  |

The 864 samples (of which 142 are healthy controls) included this study encompassed a wide range of neurologic disease across age groups, ranging from neonates (primarily IVH patients), to elderly adults with neurodegenerative disease. In all patients, the diagnosis was known, and breakdown is shown in Table 1. Of the 453 patients for whom gender was reported, 227 (50%) were male and 226 were female (50%). Of the 694 study patients in whom age was reported, 32% of patients were between 40-59 years, and 23% were between 60 and 79 years of age, age ranges where there is a high burden of neurologic disease.

### Methods of miRNA isolation and quantification in CSF

The studies comprising the super-series were selected because their approach to miRNA characterisation was unbiased, thereby facilitating the identification of biomarkers. Methods used by each study for the extraction, library preparation, and sequencing platform are summarized in Table 2, along with the number of miRNA detectable in at least 10% of samples. Most studies (7/9) relied on an Illumina platform for sequencing, one used Nanostring nCounter technology, and one used the HTG molecular diagnostics platform. Studies relying on the Illumina platform tended to use higher volumes of CSF, most commonly 1000 μL, with a yield of 47-59% of miRNA species having nonzero expression in at least 10% of samples. The HTG platform yielded a surprisingly high proportion of miRNA from 15μL of CSF, with 2066 species having non-zero reads in at least 10% of samples, though this result is based only on a single study (PREDICT-HD).^18^ Lastly, the characterization of exosomal miRNA as performed by Yagi et al., as expected, resulted in much lower yields of miRNA and required a greater quantity of CSF (7000μL).^23^

**Table 2.** Methods of RNA quantification used in studies comprising the super-series. Data from 9 studies is presented showing the variety in CSF quantity, extraction method, library preparation, sequencing method, and miRNA expressed in at least 10% of samples. N_≥10%_ refers to the proportion of quantified miRNA species with non-zero expression in at least 10% of samples. NR refers to not-reported, despite having contacted authors for study details.

| Study | CSF Quantity | Extraction | Library Preparation | Sequencing Platform | $N_{\geq 10\%}$ |
| --- | --- | --- | --- | --- | --- |
| Burgos et al., 2014 | 1000μL | mirVana<sup>TM</sup> PARIS<sup>TM</sup> kit | Illumina TruSeq Small RNA Library Prep Kit | Illumina HiSeq 2000 | 976 / 2094 (47%) |
| Dong et al., 2018 | NR | mirVana<sup>TM</sup> PARIS<sup>TM</sup> kit | Illumina TruSeq Small RNA Library Prep Kit | Illumina HiSeq 2000 | 198 / 639 (31%) |
| Drusco et al., 2015 | 250μL | Trizol extraction | nCounter Capture System | Nanostring nCounter | 225 / 225 (100%) |
| Godoy et al., 2018 | 200μL | miRNEasy Micro Kit | Illumina TruSeq Small RNA Library Prep Kit | Illumina HiSeq 4000 | 1083 / 2115 (51%) |
| Jensen, IVH | 1000μL | mirVana<sup>TM</sup> PARIS<sup>TM</sup> kit | Illumina TruSeq Small RNA Library Prep Kit | Illumina HiSeq 2000 | 1012/1705 (59%) |
| Jensen, SAH | 1000μL | mirVana<sup>TM</sup> PARIS<sup>TM</sup> kit | Illumina TruSeq Small RNA Library Prep Kit | Illumina HiSeq 2000 | 997 / 2131 (47%) |
| Reed et al., 2018 | 15μL | HTG Molecular Diagnostics miRNA whole transcriptome protocol | HTG Molecular Diagnostics miRNA whole transcriptome protocol | HTG EdgeSeq | 2066 / 2066 (100%) |
| Waller et al., 2018 | 1000μL | mirVana<sup>TM</sup> PARIS<sup>TM</sup> kit | Illumina TruSeq Small RNA Library Prep Kit | Illumina HiScanSQ | 327 / 631 (52%) |
| Yagi et al., 2017 | 7000μL (exosome sequencing) | miRNEasy Mini Kit | Illumina TruSeq Small RNA Library Prep Kit | Illumina HiSeq 2500 | 401 / 2550 (16%) |

### Broad associations of exmiRNA with categories of neurologic diseases

Expression for the 30 species of exmiRNA common to all datasets after filtering out poorly expressed species (consensus miRNA) was considered next. The expression of these 30 exmiRNA was first normalized and batch-corrected as described in the Methods. Expression was assessed for batch effects by computing the delta score (proportion of variance attributable to batch effect) and the corresponding empiric p value. Notably, pre-correction delta statistic was 0.99 with p < 0.001 for batch effect, and post-correction delta was 0.11, p = 0.99, suggesting that we had appropriately corrected for batch effects. Going forward, normalized and batch-corrected CSF exmiRNA expression was used, and a summary heatmap of normalized, batch-corrected expression for the 30 consensus exmiRNA is shown in Figure 2a. Clustering was performed using the uniform manifold approximation (UMAP) to project the expression of the consensus miRNA of each sample into two dimensions (Figure 2b), overlaid with both the diagnoses themselves, as well as the category of disease.^28^ Reproducibility of this clustering was assured with a leave-one-out approach to ensure no one dataset was skewing the results (Supplementary Figures 1-2). Hemorrhagic samples tended to cluster away from non-hemorrhagic samples, perhaps due to the miRNA intrinsic to red blood cells and platelets, and those with neurodegenerative conditions (most reliably Huntington’s disease) clustered away from healthy samples. Even despite the increased sample size, due to limitations in the number of samples for each of the conditions considered, and the heterogeneity in phenotypes, robust clustering for any particular disease based on the 30 consensus exmiRNA alone was not observed.

**Figure 2.**
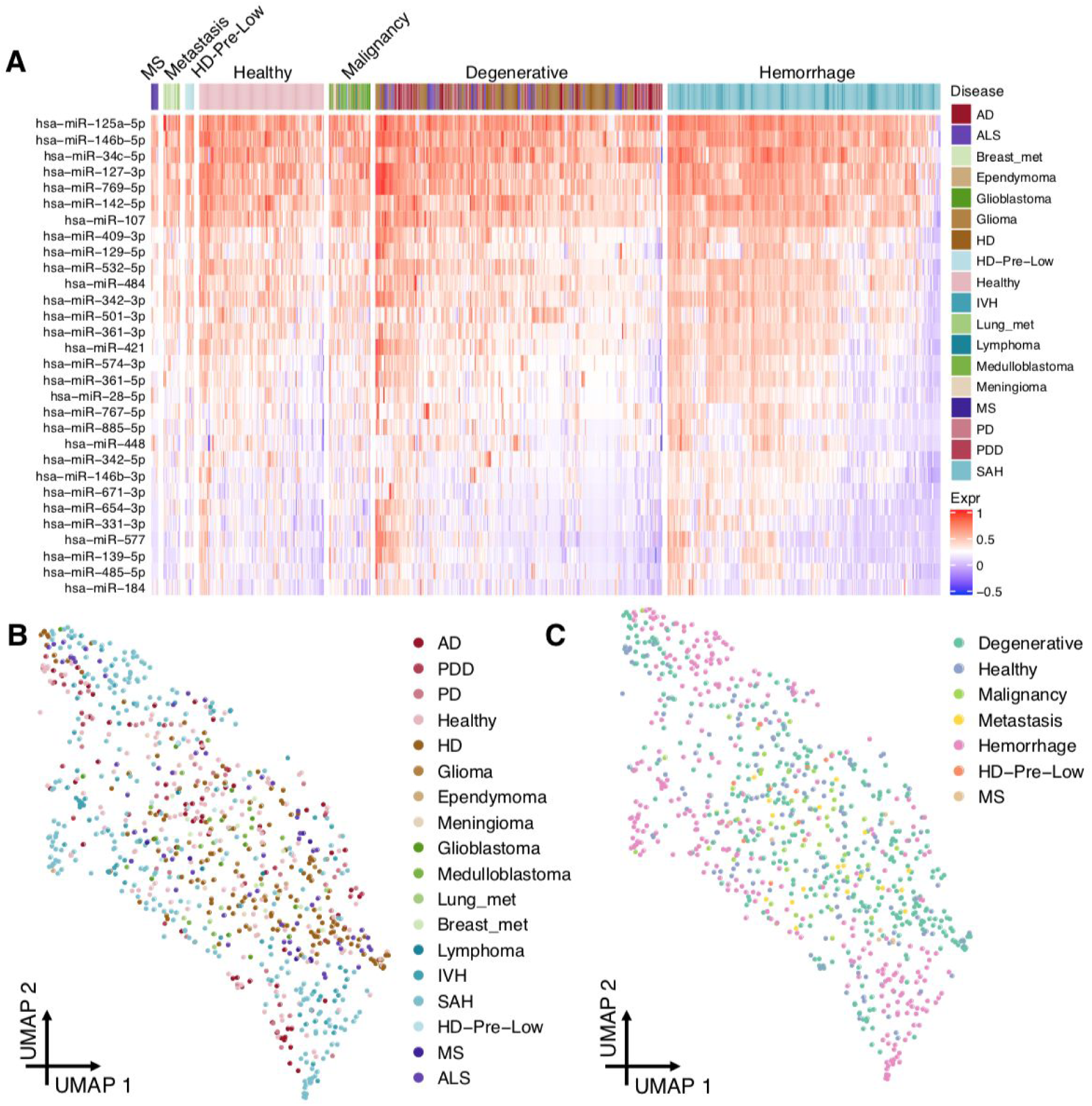
Heatmap of consensus miRNA expression and uniform manifold approximation (UMAP) plots describing sample clustering. (a) Heatmap with cells coloured by normalized batch-corrected expression for each of the 30 consensus exmiRNA identified, and samples as columns. (b) UMAP plot of samples coloured by patient condition, projecting the 30 dimensional consensus exmiRNA expression dataset into 2 dimensions, showing clustering of patients with IVH and SAH, as well as (separately), those with Huntington’s disease. (c) Samples coloured in the UMAP plot by broad category of disease as defined by disease groups (see Methods) revealing the separation between hemorrhagic CSF and degenerative CSF, and the range of exmiRNA expression in healthy patients.

### exmiRNA in healthy controls

Next, we sought to establish the range of exmiRNA expressed and detectable in the CSF of healthy controls. As such, we sought to determine which of the consensus exmiRNA showed consistently strong normalised expression in healthy samples, as assessed by the rank product statistic. 8 miRNA showed consistently strong expression above all others in healthy tissues, with statistical significance: hsa-miR-125a-5p (p < 10^-73^), hsa-miR-146b-5p (p < 10^-56^), hsa-miR-34c-5p (p < 10^-52^), hsa-miR-127-3p (p < 10^-41^), hsa-miR-142-5p (p < 10^-33^), hsa-miR-769-5p (p < 10^-31^), hsa-miR-107 (p < 10^-13^), and hsa-miR-484 (p < 0.02). Density plots of the expression of these miRNA are shown in Figure 3. Crucially, this defines a set of exmiRNA whose detectability makes them strong biomarker candidates, in which their absence or increase may signal an unhealthy state.

**Figure 3.**
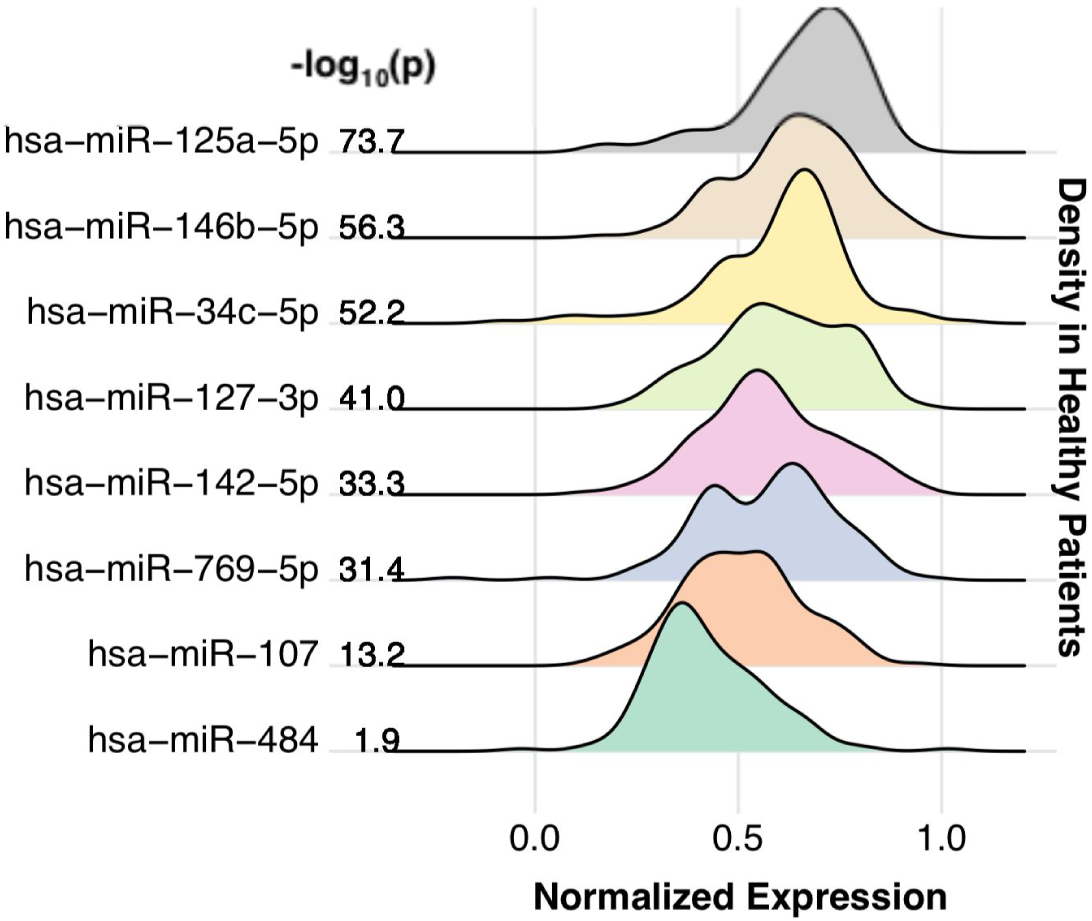
Robustly detected exmiRNA in healthy CSF. Density plots depicting expression of the 8 miRNA found to be statistically significantly detected consistently across CSF of healthy patients, as defined by the rank product. Statistical significance, p, of the rank product testing is reported as −log_10_(p). X-axis represents normalized, batch-corrected expression for each exmiRNA.

Next, we asked which of the 30 consensus miRNA could differentiate between healthy patients and those with various neurologic conditions. For each of the consensus miRNA, expression was compared in healthy patients and those with neurologic disease, to identify whether there was statistically significant difference in expression between the two states. After correcting for multiple testing, we identified four cases where miRNA expression could differentiate between the control population and a specific disease state (Figure 4a-f). Both cases identified were those of differentiating healthy patients from those with neurodegenerative conditions. In Alzheimer’s disease, hsa-miR-767-5p showed greater expression than in the control population (p < 0.001, one sided Wilcoxon rank sum test). In ALS, hsa-miR-142-5p was reduced in expression compared to the control population (p = 0.0029, one sided Wilcoxon rank sum test). In Huntington’s disease, hsa-miR-361-3p was significantly lower in expression in the HD population than control (p < 10^-4^, one sided Wilcoxon rank sum test). Similarly, in the low-risk HD premutation carriers, hsa-miR-885-5p was increased in expression compared to the control group (p = 0.0029, one sided Wilcoxon rank sum test). When considering the broad disease categories as defined previously, hsa-miR-142-5p and hsa-miR-361-3p were again identified as miRNA that tended to be reduced in expression in patients with neurodegenerative disease compared to control (p < 0.003 for both cases, one sided Wilcoxon rank sum test).

**Figure 4.**
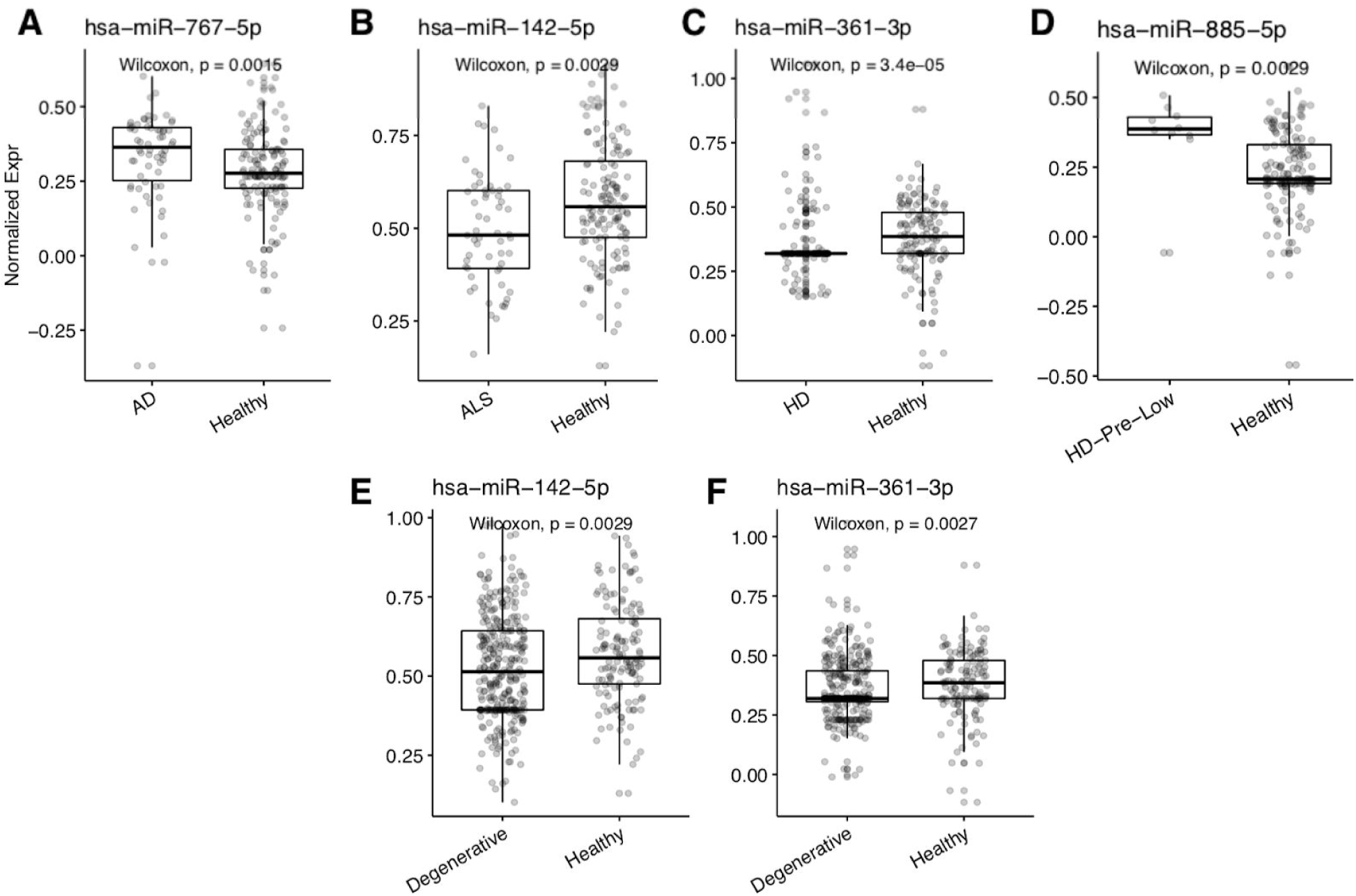
Expression of key exmiRNA differentiating disease states from healthy CSF samples. (a) hsa-miR-767-5p is statistically significantly detected at a higher level in CSF of patients with Alzheimer’s disease (AD) as compared to healthy patients. (b) hsa-miR-142-5p is detected at a lower level in CSF of patients with amyotrophic lateral sclerosis (ALS) compared to healthy controls. (c) hsa-miR-361-3p is detected at a lower level in the CSF of patients with Huntington’s disease compared to healthy controls. (d) hsa-miR-885-5p is detected at higher levels of patients with low clinical probability of Huntington’s disease (HD-Pre-Low), as defined by the PREDICT-HD study, as compared to healthy control population. (e, f) When comparing patients with degenerative conditions more generally to healthy controls, hsa-miR-142-5p and hsa-miR-361-3p show statistically significant decrease in expression.

### exmiRNA coexpression networks are dysregulated in malignancy

miRNA act as components of complex gene regulatory networks, and are expressed not in isolation, but rather in coordinated groups to alter cellular behaviour. Coexpression networks across each of the broad categories of disease as defined above were constructed, and examined for differences between diseases. Networks were defined by the Spearman correlation between different miRNA species for categories of disease in which at least 25 samples were present (healthy, degenerative, malignancy, hemorrhage), and only highly statistically significant correlations (p < 0.01) were retained for further analysis (Figure 5a). In focusing on pairs of miRNA for which there was a positive correlation in one disease subgroup and a negative correlation in another subgroup, it is evident that the underlying coexpression networks of exmiRNA are markedly different between the categories of disease considered. Among the 47 cases of miRNA-miRNA pairs that showed positive correlation in one subgroup and negative correlation in another subgroup, 45 (96%) of these involved a miRNA-miRNA pair in the malignant state. For instance, the two miRNA miR-769-5p and miR-127-3p have Spearman correlation 0.49 (p < 10^-9^) in healthy tissues, but have Spearman correlation −0.45 (p = 0.002) in malignant tissues (Figure 5b). Strikingly, this suggests that the malignant state may represent fundamentally different biology that can be detected at the level of CSF exmiRNA. Moreover, while miRNA expression within the individual samples themselves may not be expressly revealing of the malignant state, the coexpression network dysregulation compared to normal neural tissues suggests very significant underlying differences, perhaps pointing towards the use of network-based metrics as biomarkers.

**Figure 5:**
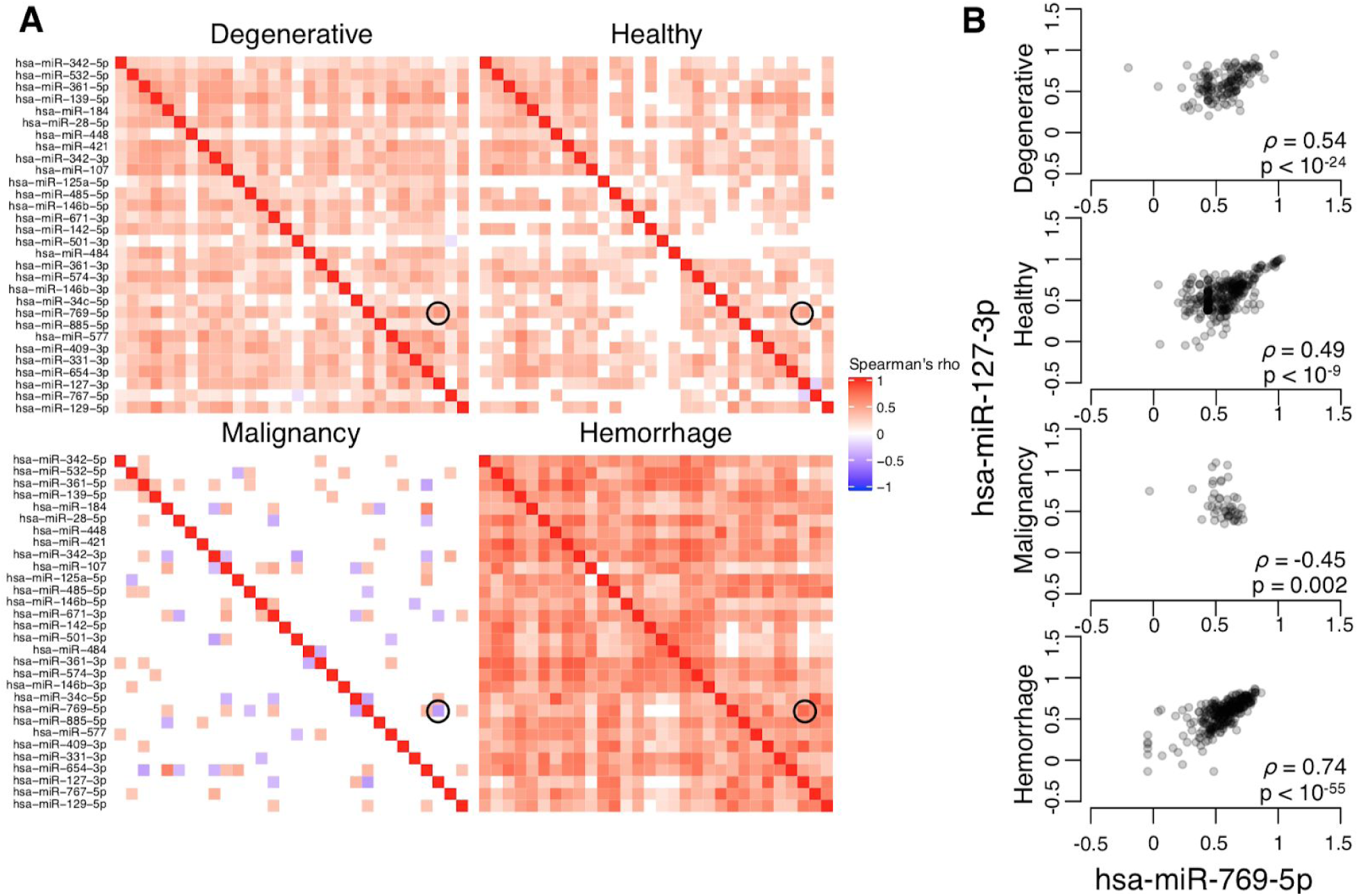
Coexpression networks in CSF exmiRNA by disease status depict the altered network in malignancy. (a) Coexpression heatmaps for consensus exmiRNA detected in the CSF of patients in degenerative, healthy control, malignancy, and hemorrhage disease categories. Values displayed are the Spearman correlation coefficient. Only statistically significant correlation values are shown (p < 0.01), and all other values are set to 0. Circled are the points on each heatmap representing the four scatterplots in (b), which show the correlation between normalized and batch corrected expression of hsa-miR-127-3p and hsa-miR-769-5p, highlighting the difference in correlation between the non-malignant and malignant states.

## Discussion

In this work, we present data collected from 9 studies that relied on unbiased sampling methods to assess exmiRNA in the CSF. This is the largest such series, and the first super-series of its kind. The datasets comprising the super-series were selected with a standardized approach, and every attempt was made to contact authors of additional relevant datasets for primary information. This study could have encompassed more smaller studies’ data, but a surprising number of authors did not respond (64%) or did not wish to share their data, or were not able to share data (36%). Despite this limitation in accruing data, this series represents a significant step forward in generating an understanding of CSF exmiRNA expression, with 864 patients of whom 142 are healthy controls, across every age group, and a wide set of 16 neurological disorders.

The identification of miRNA in biofluid is dependent on three main factors: the production and excretion of the miRNA by the cell of origin, the stability of the miRNA in the biofluid, and the technical detectability of the miRNA by extraction protocols and various sequencing technologies. Our review has shown that the majority of studies use Illumina platforms and associated protocols with 47-59% of miRNA species detectable in 90% of samples when starting with 1000μL of CSF. One study using HTG EdgeSeq had a much higher capture rate of miRNA with a significantly reduced starting volume of 15μL of CSF, suggesting that this may be a more sensitive platform, which will need to be investigated further.^18^ The role of exosomes in CSF as carrier vehicles for miRNA has shown that they are in general, less detectable than their free-floating counterparts and require a much greater quantity of CSF (7000μL) for analysis, and still have a low proportion of detectable species (16%).^23^

This super-series also provides insight into the range of exmiRNA that can be detected in the CSF of healthy patients, and it is particularly those species that are robustly detectable that may function well as biomarkers of disease. Not unexpectedly, many of the species we identified as highly expressed in CSF are known to be expressed in neural tissues themselves, such as hsa-miR-142-5p, which may function in dopaminergic signalling.^31^ hsa-miR-142-5p is also thought to contribute to pathogenesis in Alzheimer’s disease due to synaptic dysfunction, is overexpressed in patients with autism, and is downregulated in patients who completed suicide.^29–31^ Likewise, hsa-miR-107, also robustly detectable in healthy patients, has been shown to be dysregulated early in the pathogenesis of Alzheimer’s disease, with a link to increased BACE1 mRNA levels.^32,33^ hsa-miR-34c-5p has been shown to participate in cortical morphogenesis, and expression of this miRNA may be related to cognition.^34,35^ Taken together, these results suggest that the exmiNRA identified as robustly detectable in normal controls may not only represent meaningful biology, but also perhaps participate in disease pathogenesis when dysregulated, suggesting their utility as biomarkers.

Tantalizingly, our analysis also suggests that viewing miRNA as isolated variables may not reveal the full story of the biology underlying the diseased states. Rather, our work shows that the biology may be more robustly uncovered by the demonstration of altered interaction networks of miRNA. Indeed, exmiRNA coexpression networks differ significantly between malignant and non-malignant samples, suggesting network dysregulation in this state, perhaps due to a global ‘rewiring’ of the transcriptome driving disease progression and phenotype. As more datasets become available, this will be uncovered further with greater certainty, and the measurement of networks, as opposed to individual exmiRNA, may allow for greater resolution of pathophysiology.

Lastly, this dataset was not designed to be a static resource, but rather was designed with the goal of extensibility and adaptation to additional data as more becomes available. The pipeline constructed is able to facilitate comparison between any number of different datasets, while correcting for batch effects. By iteratively adding data and metadata to this resource with future studies, the field of CSF exmiRNA can be advanced faster than it would by small studies alone. It is our hope that this work can help bring insight into the instances where CSF exmiRNA biomarkers can truly alter neurology clinical practice in neurology, a field where such biomarkers are badly needed.

## Methods

### Defining the super series and literature search

The search terms used in Ovid Embase were ‘microRNA or miRNA’, ‘CSF or cerebrospinal fluid’, and ‘biomarker or bio-marker.’ Studies were limited to English language, primary research, journal articles, and human species. Only those articles with unbiased evaluation of the miRNA in the CSF (by next-generation sequencing or similar technologies), were included for further analysis. Google Scholar was used with the search term “CSF miRNA biomarker” to identify additional literature. Additional studies were obtained by searching through the Gene Expression Omnibus (GEO) database for “CSF miRNA” datasets in Homo sapiens, and in the exRNA atlas database with CSF as the primary biofluid type.^13^

### Metadata standardisation

All available metadata was acquired for each of the 9 studies included in the super-series. Across datasets, the only metadata variable that was common to all samples was diagnosis associated with the sample. Among most datasets, age of the patient at the time the sample was taken (in years), as well as the sex of the patient (male or female). These variables were collected for as many samples as possible and stored in a harmonized format. The PREDICT-HD study classified patients into one of three risk groups in addition to Huntington’s disease patients (HD-Pre-Low, HD-Pre-Med, and HD-Pre-High).^18^ For the purposes of this analysis, HD-Pre-Med and HD-Pre-High risk groups were integrated with the overall HD group and the HD-Pre-Low group was retained separately. Diagnoses were grouped into one of 5 categories for simplified analysis by broad pathophysiological mechanism. Namely, IVH and SAH were grouped into the category of hemorrhage; Alzheimer’s disease, Huntington’s disease, Parkinson’s disease, Parkinson’s disease dementia, and amyotrophic lateral sclerosis were grouped into the category of degenerative disease; glioma, ependymoma, meningioma, glioblastoma, lymphoma, and medulloblastoma were grouped into the category of malignancy; multiple sclerosis samples were maintained as multiple sclerosis, and lung and breast metastases were grouped into the category of metastasis. The remainder of the samples were healthy controls.

### Standardizing miRNA annotation version

Because different studies relied on different versions of the miRNA annotation from miRbase (ranging from v20-21), these were standardized to the latest version (v22) for the combined dataset.^24^ Crucially, because every iteration of miRbase improves upon the previous, and miR entries may be deleted as more information is obtained about various species, it was felt that it was important to harmonize before any other steps were taken, as miRNA reclassified as non-miRNA may cloud further analyses. The impact of this was relatively small, with roughly 1-2% of miRNA removed from each dataset. Most commonly, the removed species had more information in the latest version of miRbase indicating that their reads came from fragments of other larger RNA molecules, and did not represent primary miRNA on their own.

### Expression data normalisation and batch correction

All computations were done in R, version 3.5.0. Briefly, raw data were imported into R and first log-transformed using the transform log_2_(x+1) for expression x, if not already log-transformed. Subsequently, miRNA names were harmonized to version 22 of miRbase, as discussed above, using miRBaseConverter version 1.12.0.^36^ miRNA no longer considered bona fide miRNA were discarded, and in the case of non-unique mappings of miRNA after conversion, the miRNA with greater expression was retained and the others were discarded. In general, in these cases, only one of the miRNA had non-zero measurements in the original dataset. Following this, exmiRNA species with expression above 0 in at least 10% of samples were retained for further analysis. miRNA expressions were then normalized using the YuGene transform, implemented in the YuGene package version 1.1.6.^25^ After normalizing, only those miRNA common to all datasets were retained for further analysis (consensus exmiRNA). The ComBat batch correction function (as implemented in the R package sva, version 3.30.1) was then applied with batches defined as the samples from each of the 9 datasets.^37^ Batch-corrected datasets with the consensus exmiRNA were used for all further analyses. The guided principal components analysis (gPCA) package, version 1.0 was used in R to calculate delta, a test statistic estimate for the proportion of variance attributable to batch effects, along with empiric p-value, to ensure appropriate batch correction.^38^

### Association of exmiRNA to neurologic disease

The Wilcoxon rank-sum test was used to compare the batch-corrected normalized expression values across disease states, for each of the 30 consensus exmiRNAs individually. Statistical significance was defined as a one-sided Wilcoxon rank-sum test p < 0.05 after Bonferroni correction. Uniform manifold approximation (UMAP) was used to project the batch-corrected, normalized dataset into 2 dimensions using the umap package in R version 0.2.5.0.^39^ UMAP settings used were all defaults, except random seed was set to 1234567890 for reproducibility, n_neighbours was set to 30, and n_epochs was set to 2500.

### Analysis of miRNA expressed in healthy tissues

To obtain the miRNA most consistently showing strong expression in healthy samples, the batch-corrected normalized dataset was subsetted to include only healthy samples, across the 30 consensus miRNA. Following this, across samples, the rank product statistic was used to identify the miRNA species that consistently ranked high in expression across the healthy samples. The rank product test statistic and empiric p-value were computed using base settings and functions from the RankProd package, version 3.14.0 in R.^40^ Statistical significance was defined as Bonferroni corrected p < 0.05.

### exmiRNA coexpression network

Coexpression network of miRNA was constructed by computing the Spearman correlation between pairs of the consensus exmiRNA in groups of samples as stratified by their disease category. To ensure that correlations observed were above a minimum threshold of statistical significance, only disease categories with at least 25 samples were used. Correlation coefficient was defined as Spearman’s rho if the value was statistically significant (p < 0.01), and was 0 otherwise.

## Dataset and code availability

The final dataset cleaned with normalised, batch-corrected expression values has been uploaded to Github (https://github.com/andrewdhawan/csf_ncRNA) as a.RDA file, along with all code that reproduces the plots and analysis within this paper.

## Acknowledgements

A.D. is indebted to not only the authors of all of the studies cited whose data was able to be compiled into this immense resource, but also to the patients and families themselves offering CSF for study purposes.

## Authors’ Contributions

A.D. designed the study, performed the analyses, and wrote the manuscript.

## Ethics Approval

Study design was to use deidentified patient data already in the published domain, and as such, ethics approval was not required. All original data were collected with requisite ethics approval as cited in the primary manuscripts.

## Supplementary Information

**Supplementary Figure 1.**
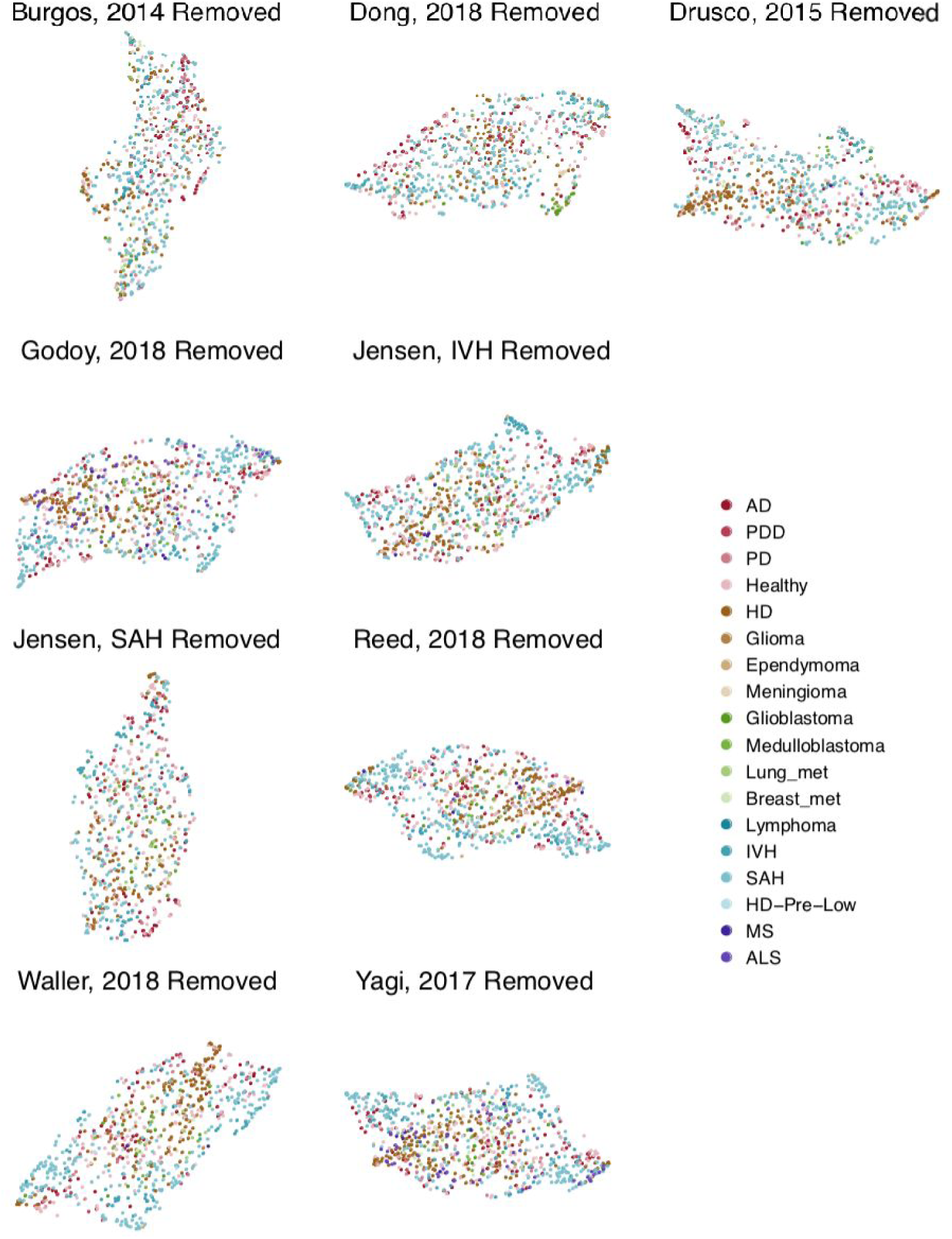
UMAP plots coloured by disease for leave-one-out dataset. Data was pre-processed and batch corrected with the indicated dataset removed. Clustering was then performed using the resultant consensus exmiRNA common to all datasets included. UMAP plots were then regenerated (n_neighbours = 30, n_epochs = 2500) and show similar clustering as the full super-series, suggesting that no one dataset significantly skewed UMAP results.

**Supplementary Figure 2.**
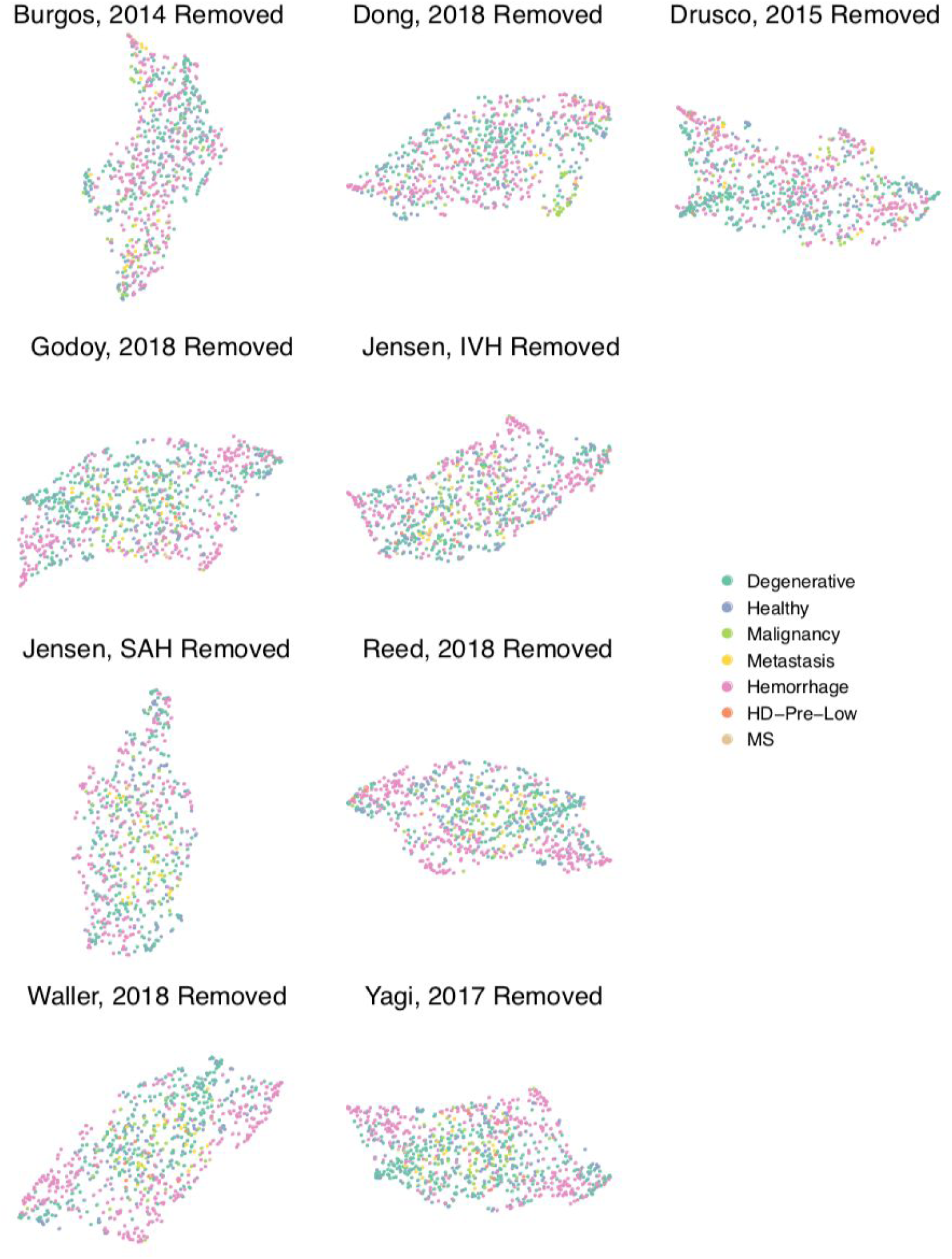
UMAP plots coloured by disease category for leave-one-out datasets. As in Supplementary Figure 1, data were pre-processed and batch-corrected with the indicated dataset removed. As in the full super-series, exmiRNA were again restricted to those common between all datasets included, and clustering with UMAP was performed in two dimensions (n_neighbours = 30, n_epochs = 2500), and points coloured by disease category. Again, similar clustering as in the full super-series is observed, suggesting that there is no strong driving influence by any one dataset in the UMAP analysis.

